# Integrative Omics Analysis Reveals Functional Variants of Hemoglobin Zeta (HBZ) as an Ancestry-Specific Risk Factor for Triple-Negative Breast Cancer

**DOI:** 10.64898/2026.10.08.26364913

**Authors:** Han Le, Amos Sakwe, Ashton Oliver, Ayo P. Doumatey, Ky’Era Actkins, Mileati Melese, Rokhaya Ndiaye, Amadou Gaye¹

## Abstract

**Background:** Triple-negative breast cancer (TNBC) disproportionately affects women of African ancestry, yet the biological mechanisms underlying this disparity remain poorly understood. While recent genome-wide association studies (GWAS) have identified numerous TNBC risk variants, their functional consequences have not been systematically evaluated in African-ancestry populations. We hypothesized that ancestry-specific regulatory variants drive distinct transcriptomic and proteomic landscapes that contribute to TNBC aggressiveness.

**Methods:** We extracted 2,849 unique TNBC-associated variants (p-value ≤ 1×10□□) from the GWAS Catalog. Of these, 206 were excluded due to absence in our whole-genome sequencing (WGS) dataset or low minor allele frequency (<0.01). We prioritized the remaining 2,643 bi-allelic variants for analysis in the GENE-FORECAST cohort. We integrated WGS with whole-blood transcriptomic (n=242) and proteomic data (n=362) to perform cis-eQTL and cis-pQTL analyses in discovery and validation sets.

**Results:** We identified 38 cis-eQTL associations and 5 cis-pQTL associations that were statistically replicated across independent discovery and validation datasets within our cohort. Both omics layers converged on a single gene, Hemoglobin Zeta (*HBZ*). We identified five functional variants (four SNPs and one INDEL) in strong linkage disequilibrium that consistently upregulate *HBZ* mRNA and the corresponding circulating embryonic zeta-globin protein. These variants were found almost exclusively in African and African-admixed populations across GENE-FORECAST, 1000 Genomes, BioVU, and All of Us datasets. Four of the five variants were also confirmed as significant HBZ eQTLs in GTEx whole blood.

**Conclusion:** Our integrative analysis reveals variants of the embryonic *HBZ* gene as a novel, ancestry-specific TNBC-associated risk factor that is detectable at both transcriptomic and proteomic levels. This suggests that aberrant reactivation of this embryonic globin may provide a selective advantage in the hypoxic TNBC microenvironment. These results highlight a unique mechanism of tumorigenesis in women of African ancestry, offering a potential distinct biomarker and therapeutic target for this high-risk population.

Proposed Model of HBZ-Driven TNBC Aggressiveness in African Ancestry. Schematic illustrating the hypothesized biological mechanism linking genetic ancestry to TNBC disparity. (Left) Ancestry-specific regulatory variants located at the 16p13.3 locus are inherited predominantly in individuals of African descent. (Middle) In normal adult tissues, the embryonic HBZ gene is canonically silenced. However, the presence of these risk variants disrupts regulatory elements, leading to the aberrant reactivation and ectopic expression of HBZ mRNA and protein in breast tissue. (Right) We hypothesize that the re-expression of this embryonic hemoglobin provides TNBC cells with a unique oxygen-scavenging advantage, enabling survival in hypoxic tumor microenvironments, promoting aggressiveness, and contributing to poorer clinical outcomes observed in women of African ancestry.

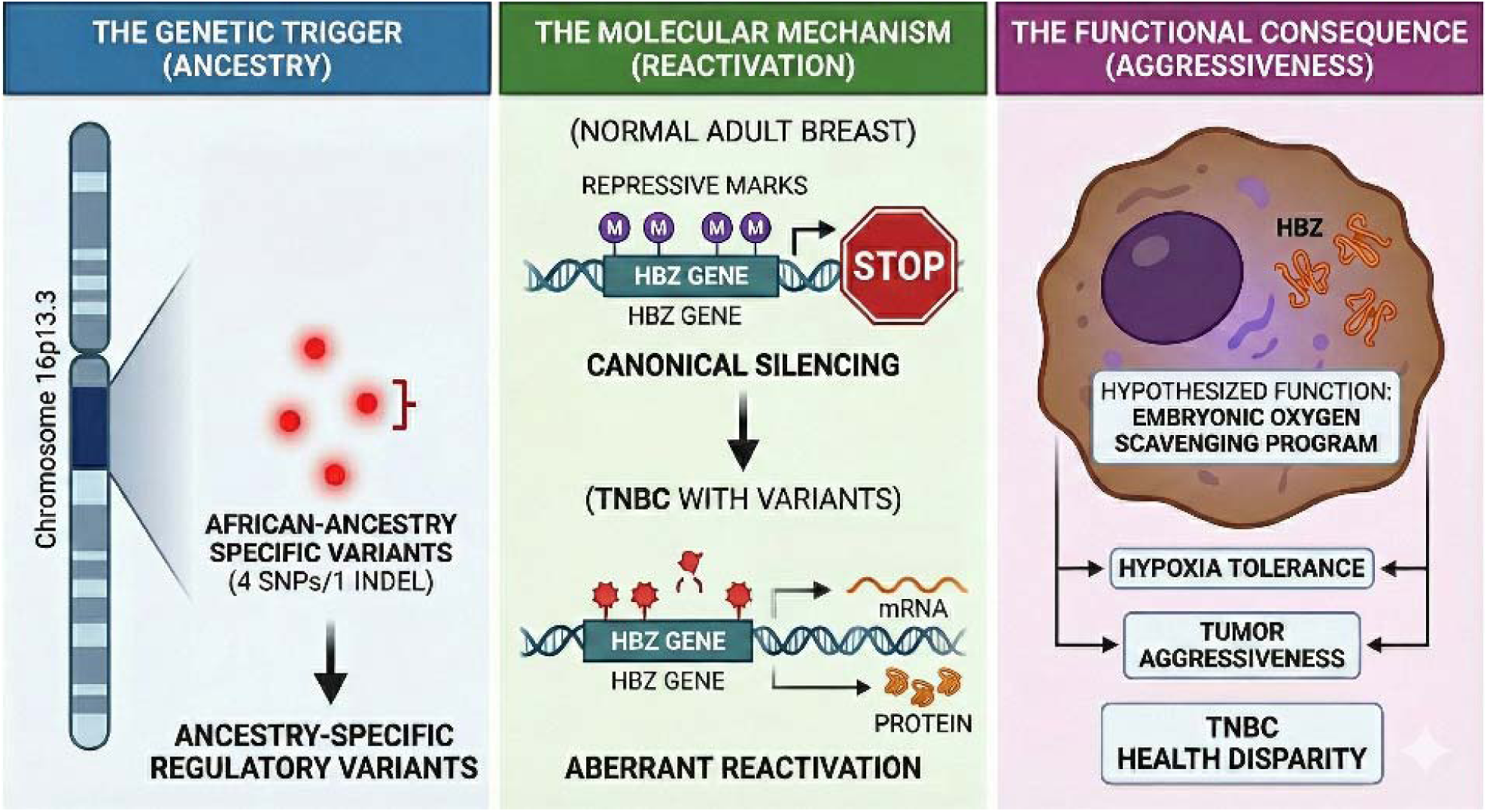

## INTRODUCTION

Triple-Negative Breast Cancer (TNBC) is an aggressive and clinically challenging subtype of breast cancer ^1^. It is characterized by the lack of expression of the estrogen receptor (ER), progesterone receptor (PR), and human epidermal growth factor 2 (HER2) ^2^. This absence of key receptors renders it unresponsive to standard of care hormonal or HER2-targeted therapies, leaving systemic chemotherapy as the primary treatment modality^3^. It is associated with a poor prognosis, exhibiting a tendency for early recurrence and an increased risk of metastasis ^3^. Biologically, its rapid and chaotic cell proliferation often outpaces the development of an efficient blood supply, creating a severely low-oxygen (hypoxic) tumor microenvironment, which itself is a known driver of tumor progression and therapeutic resistance ^4^.

A significant and persistent health disparity exists, as TNBC disproportionately affects women of African, African American, and Latin American descent ^5, 6^. TNBC accounts for approximately 10-20% of breast-cancer diagnoses in high-income Western populations ^1^, but the proportion is substantially higher in West Africa, where estimates range from ∼40% to 50% ^7^. This striking difference in prevalence strongly suggests that genetic factors contribute to TNBC susceptibility, although socio-environmental determinants and structural inequities undoubtedly play important roles. Given the heterogeneity of TNBC, a multi-omic approach to tumor characterization is essential to deconstruct these variables. Yet progress remains limited by the persistent underrepresentation of African American and African-ancestry women in large-scale cancer genomic studies, despite their disproportionate disease burden.

Large-scale Genome-Wide Association Studies (GWAS) ^8^ have been successful in identifying numerous genetic loci associated with breast cancer risk. These studies have significantly expanded the map by identifying new susceptibility loci through both overall and subtype-specific analyses ^9^. Other work has specifically confirmed that many known breast cancer loci are also significant risk factors for the TNBC subtype ^10^. More recently, research has focused on the genetic differences between subtypes and in diverse populations. One study used a case-by-case method to directly compare breast cancer subtypes, successfully identifying genetic variants that specifically inform on risk for TNBC versus luminal breast cancer subtypes ^11^. Most pertinent to the health disparity, a recent GWAS focused exclusively on women of African ancestry not only identified new susceptibility loci for this population but also demonstrated that genetic risk models built from this data are more predictive than those based on European-ancestry cohorts ^12^.

However, linking these statistical associations to a functional biological consequence remains a primary challenge. Most of these variants lie in non-coding regions of the genome, and their effects on gene regulation are largely unknown. This gap is particularly wide in non-European populations, as most genomic studies have focused on European-ancestry cohorts, hindering our understanding of the specific mechanisms that may be driving TNBC-associated health disparities ^12^.

The objective of this study was to bridge this gap by integrating GWAS data with an unbiased, integrative multi-omics analysis to identify novel genes that could explain TNBC disparities. We leveraged data from the GENE-FORECAST (Genomics, Environmental Factors, and the Social Determinants of Cardiovascular Disease in African Americans Study) cohort ^13^, which includes self-identified African Americans, to agnostically screen for the functional consequences of known TNBC-associated variants. By integrating Whole-Genome Sequencing (WGS) data with transcriptomic (mRNA-seq) and proteomic (Olink) data, we performed expression (eQTL) and protein (pQTL) quantitative trait loci analyses. A key strength of this approach was the concurrent use of two distinct high-throughput technologies to analyze the genetic effect on both mRNA and protein from the same whole blood samples within the cohort. The convergence of multi-omics data was used to identify and prioritize the most relevant, functionally supported candidate genes underlying this disparity without imposing any pre-specified targets.

## MATERIALS AND METHODS

### Study Cohort

GENE-FORECAST is a research platform strategically designed to employ a comprehensive, multi-omics systems biology approach for in-depth, multi-dimensional phenotyping of health and disease within the African American population. Utilizing a community-based sampling framework, GENE-FORECAST established a cohort comprising U.S.-born African American men and women aged 21-65, recruited primarily from the metropolitan Washington D.C. area as described previously ^13^. GENE-FORECAST was used as approved by the Institutional Review Boards (IRB) of the National Institutes of Health and Meharry Medical College. All participants provided written informed consent prior to their involvement. The project presented here was conducted in compliance with local regulations and institutional protocols.

### Genotype Data

The genotype data are from WGS. DNA was extracted from whole blood collected in ethylenediaminetetraacetic acid (EDTA) coated tubes, followed by picoGreen quantitation. Library preparation was done using Whole Genome Small Insert Polymerase Chain Reaction (PCR)-Free. The WGS samples were 151bp paired-end sequenced on an Illumina NovaSeq6000 to a mean coverage of 30X. At the data preprocessing step, WGS reads were processed with the Whole Genome Germline Variant Discovery pipeline developed and used by the Genomics Platform at the Broad Institute. Reads were aligned to the genome build Hg38 with Burrows-Wheeler Aligner, followed by Genomic Variant Call Format (gVCF) generation, joint genotype calling, and quality filtering executed in accordance with Genome Analysis Toolkit (GATK4) best practices. Of the 371 samples with whole-genome sequencing (WGS) data, 242 also had whole-blood gene expression profiles obtained through mRNA sequencing, and 362 had plasma protein levels measured using the Olink Explore 3072 platform.

### Transcriptomics Data

The transcriptome data consists of the messenger RNA sequencing (mRNA-seq) from 497 whole blood samples. Total RNA extraction was carried out using MagMAX^TM^ for Stabilized Blood Tubes RNA Isolation Kit (Life Technologies, Carlsbad, CA). For the library preparation, total RNA samples were converted into indexed cDNA sequencing libraries using Illumina’s TrueSeq kits. Ribosomal RNA (rRNA) was removed. The GENE-FORECAST samples were pair-end sequenced on the Illumina HiSeq2500 and HiSeq4000 platforms with a sequencing depth of at least 50 million reads per sample. To measure gene expression levels, mRNA expression was quantified using a bioinformatics pipeline developed by the Broad Institute and used by the Genotype-Tissue Expression (GTEx). The pipeline is detailed in the GitHub software development platform ^14^. Transcripts with expression levels below 2 counts per million (CPM) in at least 90% of samples were excluded from the analysis. The expression data were normalized using the Trimmed Mean of M-values (TMM), an optimal method for read count data ^15^. Principal component analysis (PCA) was conducted to identify and exclude transcript outliers; 4 transcripts were removed. A total of 129,921 mRNA transcripts from 16,247 protein-coding genes passed quality control and were included in the expression quantitative trait loci (eQTL) analysis, conducted in the subset of 242 samples with available WGS data.

### Proteomics Data

The Explore 3072 assay, utilizing 8 Explore 384 Olink panels (Cardiometabolic, Cardiometabolic II, Inflammation, Inflammation II, Neurology, Neurology II, Oncology, Oncology II), was used to assess the relative expression of a total of 2,947 proteins. Proximity Extension Assay (PEA) technology was conducted according to the Olink AB manufacturer’s procedures by the certified laboratory. Briefly, the technique relies on the use of antibodies labelled with unique DNA oligonucleotides that bind to their target protein present in the sample. The DNA oligonucleotides, when in proximity of a target protein, undergo hybridization and act as a template for DNA polymerase-dependent extension, forming a unique double-stranded DNA barcode proportionate to the initial protein concentration. Quantification of resulting DNA amplicons is accomplished through high-throughput DNA sequencing, generating a digital signal reflective of the number of DNA hybridization events corresponding to the protein concentration in the original sample. The measurement of protein levels is based on Normalized Protein eXpression (NPX) values, serving as a relative protein quantification unit. This quantification is normalized to account for systematic noise arising from sample processing and technical variation, leveraging internal controls and sample controls. NPX units are on a log2 scale, where one NPX unit increase indicates a two-fold rise in the concentration of amplicons representing the target protein compared to the internal control. A total of 2,941 proteins that passed vendor quality-control checks were included in the protein quantitative trait loci (pQTL) analysis, which was performed using the 362 samples with available WGS data.

### Statistical Analyses

The GENE-FORECAST data were randomly split into two datasets: the discovery dataset (n = 121 for eQTL; n = 181 for pQTL) and the validation dataset (n = 121 for eQTL; n = 181 for pQTL). The analyses were conducted in three main steps, depicted graphically in Figure 1. First, summaries from GWAS studies of TNBC deposited in the GWAS catalog were downloaded. Genetic variants reported to be associated with TNBC at a p-value ≤ 0.0001 were extracted. The decision to extract variants associated with TNBC at a nominal ≤ 0.0001, a threshold below the genome-wide significance standard, was based on the recognition that variants below this conventional statistical threshold may still significantly impact gene expression or protein function. This approach is justified by the growing biological evidence supporting the functional relevance of these sub-threshold variants, making them essential candidates for integrative multi-omics analysis ^16–19^.

**Figure 1:**
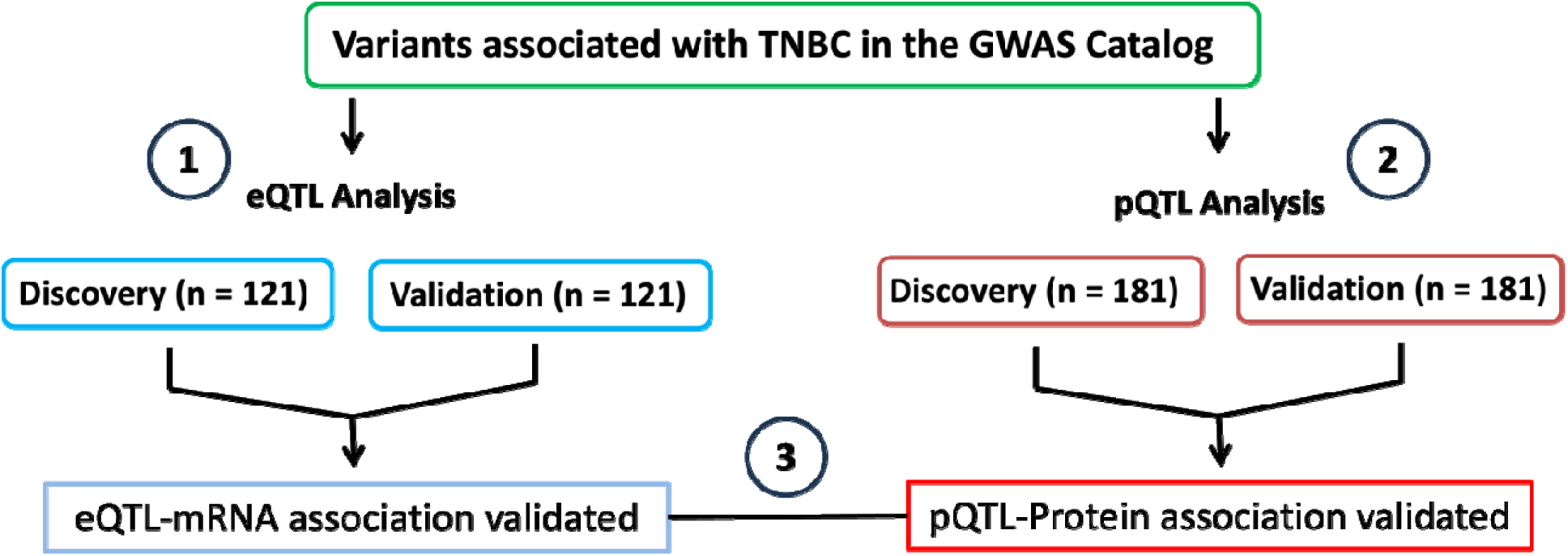
Integrative Multi-Omics Workflow. Schematic overview of the analytical pipeline used to identif functional TNBC risk variants. The GENE-FORECAST cohort was randomly split into independent Discovery and Validation datasets. (1) eQTL Analysis: We tested 2,643 TNBC-associated variants for association with mRNA expression (n=242). (2) pQTL Analysis: The same variants were tested for association with plasma protein levels (n=362). (3) Integration: We prioritized candidates that showed replicated, concordant effects across both transcriptomic and proteomic layers. Only variants passing FDR correction in the discovery set and nominal significance in the validation set were retained.

Then, eQTL and pQTL analyses were conducted with that set of SNPs. Finally, the relationship between genes and proteins linked to the eQTL and pQTL variants was investigated using the whole GENE-FORECAST dataset.

eQTL and pQTL analyses were performed to assess the impact of genetic variants previously associated with TNBC in GWAS on gene expression and protein abundance, thereby providing insights into the molecular mechanisms linking GWAS-identified variants to TNBC.

Both eQTL and pQTL analyses were conducted utilizing the R library *MatrixEQTL* ^20^. *MatrixEQTL* applies a regression model with mRNA/protein levels as the outcome and additive genotypes as independent variables. The regression model was adjusted for covariates, including age, sex, and Principal Components (PCs) 1 to 6 that explain most of the variance, to account for genetic ancestry admixture. The analyses were restricted to variants located within the cis region, spanning 1 megabase of the mRNA for eQTL or the mRNA encoding the protein for pQTL.

A variant was designated as an eQTL if it met all three criteria: (a) the False Discovery Rate (FDR)-adjusted p-value in the discovery set was equal to or less than 0.05; (b) the nominal p-value in the validation dataset is equal to or less than 0.05; and (c) the beta value of the association of the variant with an mRNA was in the same direction in the discovery and validation datasets. Analogously, a variant was deemed a pQTL if the beta value and the p-value of the association with a protein met the same three criteria.

The focus on bi-allelic variants for the analyses was grounded on multiple considerations, including the higher accuracy and reliability associated with genotyping techniques for bi-allelic SNPs compared to multi-allelic variants. This increased precision contributes to the robustness of our pQTL and eQTL findings. Furthermore, the choice of bi-allelic variants is strategically driven by their binary allele composition, facilitating the interpretation of genetic effects and streamlining the identification and characterization of associations between specific alleles and mRNA and protein levels.

All statistical analyses included both male and female participants. Although breast cancer predominantly affects women, it also occurs in men, representing a small but clinically meaningful subset of male breast cancers ^21^. In addition, the National Institutes of Health policy requires the inclusion of sex as a biological variable in human studies unless a strong scientific rationale exists for exclusion ^22^, which is not the case for breast cancer. Therefore, incorporating both male and female participants enhances rigor, generalizability, and the interpretability of molecular findings.

## RESULTS

### Data Analysis

We analyzed 242 samples for the eQTL analysis and 362 samples for the pQTL analysis, each divided evenly into discovery and validation datasets (eQTL: 121/121; pQTL: 181/181). **Table 1** summarizes the demographic and clinical characteristics of participants, including age, sex, systolic and diastolic blood pressure, kidney function (eGFR), anthropometric measures, lipid parameters, and current smoking status. Across both the eQTL and pQTL analyses, there were no statistically significant differences between the discovery and validation cohorts for any variable. This balance indicates that the datasets are well-matched and suitable for the discovery and replication of QTL signals.

**Table 1:**
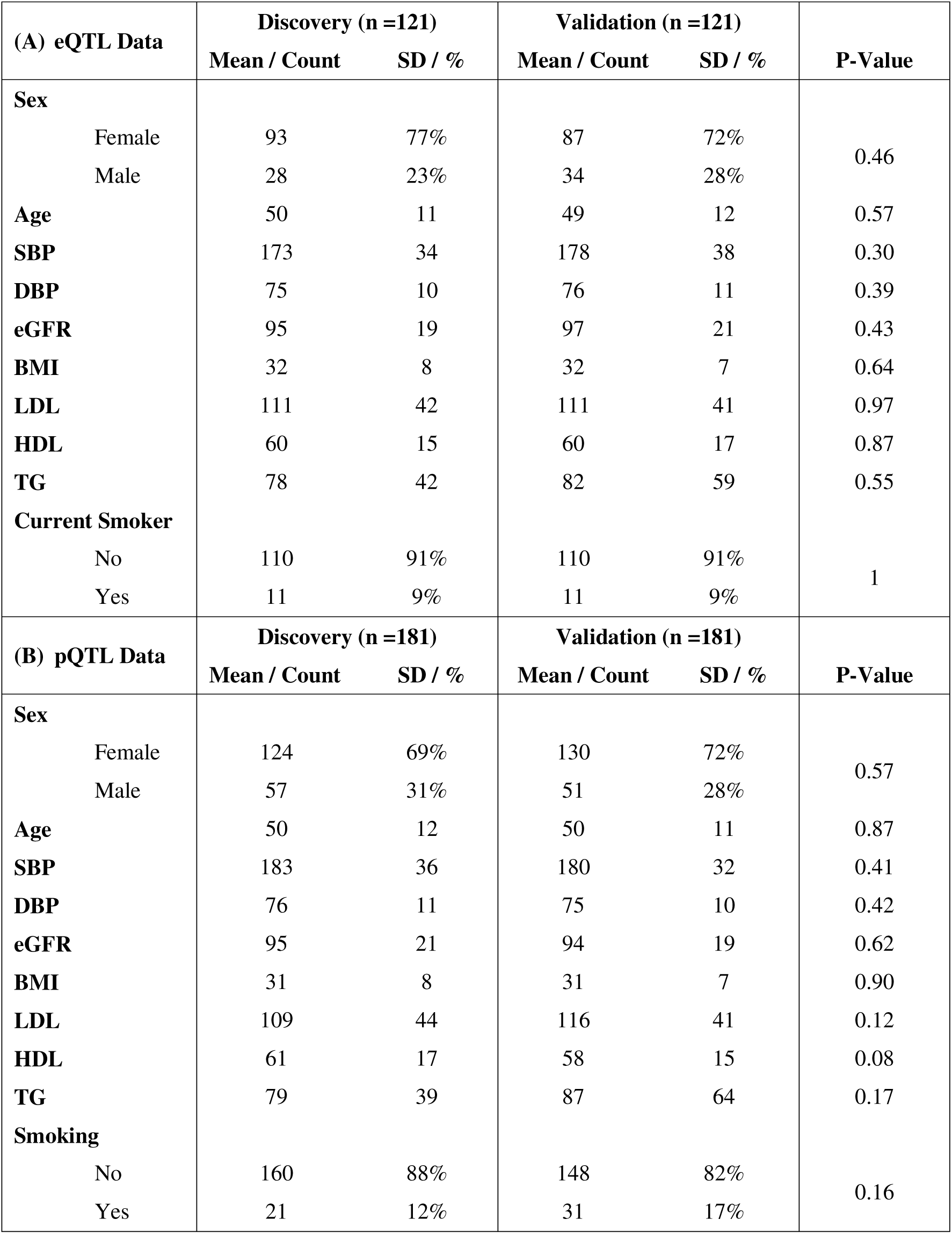
Baseline characteristics of samples included in the eQTL and pQTL analyses.

### GWAS-Derived TNBC Risk Loci Used for QTL Analyses

Using the GWAS Catalog Trait ID for triple-negative breast cancer (EFO_0005537), we identified four TNBC GWAS published between 2020 and 2024. We downloaded the corresponding summary statistics and extracted all variants with an association p-value ≤ 1×10□□ for downstream analyses. The four GWAS included in this study are listed in **Table 2**.

**Table 2:** TNBC genome-wide association studies (GWAS) retrieved from the GWAS Catalog.

| GWAS Catalog Study Accession | PubMed ID | Publication Date (Author) | Reported Trait | Study Sample Description |
| --- | --- | --- | --- | --- |
| GCST90296722 | 38741014 | 2024-05-13 (Jia et al.) | Breast cancer (estrogen-receptor negative, progesterone-receptor negative, and human epidermal growth factor-receptor negative) | 2,860 African or African American cases<br>16,262 African or African American controls |
| GCST002305 | 24325915 | 2013-12-09 (Purrington et al.) | Breast cancer (estrogen-receptor negative, progesterone-receptor negative, and human epidermal growth factor-receptor negative) | 1,529 European ancestry cases<br>3,399 European ancestry controls |
| GCST90446468-74 | 38832928 | 2024-06-04 (Sun et al.) | Breast cancer subtype (triple negative vs luminal A-like) | 16,499 European ancestry triple negative cases,<br>60,715 European ancestry luminal A-like cases |
| GCST90454344 | 32424353 | 2020-05-18 (Zhang et al.) | Breast cancer (triple negative) | ~14,900 European ancestry cases<br>91,477 European ancestry controls |

We used genomic coordinates based on the Genome Reference Consortium Human Build 38 (GRCh38) to intersect the GWAS-identified variants with bi-allelic variants present in the GENE-FORECAST whole-genome sequencing dataset, restricting to variants with a minor allele frequency (MAF) ≥ 0.01. Of the variants extracted from the TNBC GWAS summary statistics, 34,176 bi-allelic SNPs were found in the GENE-FORECAST WGS dataset.

Across the four TNBC GWAS reports examined, a total of 2,849 unique variants were documented in the GWAS Catalog. Of these, 2,783 variants (97.7%) were successfully captured in our whole-genome sequencing dataset. After applying a minor allele frequency threshold of MAF ≥ 0.01, 2,643 variants were retained for downstream analyses (**Table 3**).

**Table 3:** Recovery of TNBC GWAS variants in the GENE-FORECAST whole-genome sequencing dataset.

| Author | PubMed ID | # in GWAS Catalog | # in our WGS | # with MAF $\geq 0.01$ |
| --- | --- | --- | --- | --- |
| Jia G | 38741014 | 2816 | 2752 | 2613 |
| Purrington KS | 24325915 | 3 | 3 | 3 |
| Sun X | 38832928 | 27 | 26 | 25 |
| Zhang H | 32424353 | 3 | 2 | 2 |
| Total | - | 2849 | 2783 | **2643** |

The largest contribution by variant number came from Jia et al., which reported 2,816 variants, of which 2,752 were identified in our WGS data and 2,613 met the MAF threshold. By contrast, Purrington et al., Sun et al., and Zhang et al. reported only a small number of loci. This is because these studies did not deposit full summary statistics and instead listed only genome-wide significant variants. As a result, their contributions to the variant pool were relatively small; however, all reported variants were nearly fully recovered in our WGS dataset.

### eQTL identified 24 unique variants from18 genes

The eQTL analysis was conducted using 129,921 mRNA transcripts from 16,247 protein-coding genes. Associations between the 2,643 GWAS genetic variants and transcript expression were first assessed in a discovery set of 121 samples with WGS data and then evaluated in a replication set of 121 samples. Using a joint significance criterion of an FDR-adjusted p-value ≤ 0.05 in one dataset and a nominal p-value ≤ 0.05 in the other dataset, we validated 38 eQTL-mRNA associations, involving 24 unique variants and 21 unique mRNA transcripts from 18 genes (**Table 4**).

**Table 4:** Top replicated cis-eQTLs ranked by significance in the discovery dataset. Each of the variants reached a GWAS p-value ≤ 0.0001 in one or more of the TNBC GWAS studies listed in Table 3. For each eQTL, genomic position in the GRCh38 build, target gene/transcript, and effect estimates (β), raw p-values, and FDR-adjusted p-values in the discovery set are shown, along with corresponding β and p-values in the validation set. All associations shown met replication criteria (FDR ≤ 0.05 in discovery and nominal p ≤ 0.05 in validation).

| eQTL | Chr. | Position (Build 38) | Gene | Discovery |  |  | Validation |  |
| --- | --- | --- | --- | --- | --- | --- | --- | --- |
|  |  |  |  | Beta | P | FDR | Beta | P |
| rs111760662 | 11 | 93868749 | *C11orf54* | 405.35 | 1.20e-13 | 1.27e-09 | 120.79 | 3.66e-03 |
| rs111401516 | 11 | 93897410 | *C11orf54* | 405.35 | 1.20e-13 | 1.27e-09 | 120.79 | 3.66e-03 |
| rs111760662 | 11 | 93868749 | *ENST00000638767* | 189.10 | 1.19e-11 | 7.81e-08 | 59.30 | 7.10e-04 |
| rs111401516 | 11 | 93897410 | *ENST00000638767* | 189.10 | 1.19e-11 | 7.81e-08 | 59.30 | 7.10e-04 |
| rs111760662 | 11 | 93868749 | *C11orf54* | 277.46 | 1.93e-11 | 1.12e-07 | 106.83 | 9.33e-03 |
| rs111401516 | 11 | 93897410 | *C11orf54* | 277.46 | 1.93e-11 | 1.12e-07 | 106.83 | 9.33e-03 |
| rs75068884 | 13 | 37648916 | *POSTN* | 0.78 | 1.59e-08 | 3.59e-05 | 0.09 | 2.28e-03 |
| rs13336827 | 16 | 143089 | *HBZ* | 113.38 | 4.97e-08 | 7.83e-05 | 43.44 | 6.69e-03 |
| rs149127794 | 16 | 146167 | *HBZ* | 106.97 | 1.24e-07 | 1.70e-04 | 37.59 | 1.68e-02 |
| rs13337843 | 16 | 156204 | *HBZ* | 106.00 | 1.66e-07 | 2.21e-04 | 50.27 | 8.02e-04 |
| rs13331856 | 16 | 156229 | *HBZ* | 105.80 | 1.78e-07 | 2.33e-04 | 50.27 | 8.02e-04 |
| rs116729640 | 13 | 37623994 | *POSTN* | 0.65 | 3.88e-07 | 4.36e-04 | 0.09 | 2.28e-03 |
| rs112234623 | 16 | 157048 | *SNRNP25* | 2.72 | 1.48e-06 | 1.28e-03 | 1.62 | 1.01e-02 |
| rs149127794 | 16 | 146167 | *SNRNP25* | 2.76 | 2.07e-06 | 1.74e-03 | 1.87 | 3.58e-03 |
| rs13337843 | 16 | 156204 | *SNRNP25* | 2.73 | 2.91e-06 | 2.25e-03 | 1.71 | 5.84e-03 |
| rs141732375 | 19 | 51341816 | *ZNF766* | 6.60 | 2.99e-06 | 2.29e-03 | 3.34 | 7.95e-03 |
| rs78488761 | 19 | 51331131 | *ZNF766* | 6.60 | 2.99e-06 | 2.29e-03 | 3.34 | 7.95e-03 |
| rs183886108 | 19 | 51328214 | *ZNF766* | 6.60 | 2.99e-06 | 2.29e-03 | 3.34 | 7.95e-03 |
| rs13331856 | 16 | 156229 | *SNRNP25* | 2.72 | 3.12e-06 | 2.37e-03 | 1.71 | 5.84e-03 |
| rs112234623 | 16 | 157048 | *HBZ* | 90.67 | 5.41e-06 | 3.95e-03 | 40.75 | 7.85e-03 |
| rs34673822 | 17 | 58272008 | *MKS1* | 5.88 | 1.14e-05 | 7.27e-03 | 2.43 | 3.57e-02 |
| rs72838029 | 2 | 117989583 | *DDX18* | 16.09 | 1.78e-05 | 1.06e-02 | 4.08 | 3.86e-03 |
| rs371006416 | 1 | 38651104 | *MANEAL* | 14.55 | 2.57e-05 | 1.46e-02 | 10.20 | 1.41e-02 |
| rs12879411 | 14 | 23208960 | *RBM23* | 54.44 | 2.76e-05 | 1.55e-02 | 70.94 | 1.57e-04 |
| rs149127794 | 16 | 146167 | *SNRNP25* | 7.83 | 4.05e-05 | 1.90e-02 | 8.75 | 1.09e-04 |
| rs13331856 | 16 | 156229 | *SNRNP25* | 7.80 | 4.30e-05 | 1.96e-02 | 8.64 | 7.52e-05 |
| rs111752552 | 1 | 3114551 | *RER1* | 102.46 | 4.32e-05 | 1.97e-02 | 62.92 | 4.69e-03 |
| rs13337843 | 16 | 156204 | *SNRNP25* | 7.79 | 4.38e-05 | 2.00e-02 | 8.64 | 7.52e-05 |
| rs114609141 | 15 | 34114519 | *EMC4* | 5.72 | 4.41e-05 | 2.00e-02 | 2.52 | 2.83e-03 |
| rs12434743 | 14 | 104210771 | *MARK3* | 4.06 | 6.34e-05 | 2.60e-02 | 1.83 | 2.93e-03 |
| rs111760662 | 11 | 93868749 | *C11orf54* | 954.91 | 7.37e-05 | 2.88e-02 | 637.15 | 4.95e-04 |
| rs111401516 | 11 | 93897410 | *C11orf54* | 954.91 | 7.37e-05 | 2.88e-02 | 637.15 | 4.95e-04 |
| rs13336827 | 16 | 143089 | *SNRNP25* | 2.42 | 7.55e-05 | 2.92e-02 | 2.03 | 1.97e-03 |
| rs7189896 | 16 | 83241417 | *ADAD2* | 0.58 | 9.36e-05 | 3.43e-02 | 0.40 | 1.31e-03 |
| rs113129808 | 5 | 429467 | *EXOC3* | 88.12 | 1.34e-04 | 4.35e-02 | 107.56 | 3.55e-06 |
| rs73819804 | 5 | 160302450 | *SLU7* | 1.96 | 1.36e-04 | 4.35e-02 | 0.21 | 2.07e-03 |
| rs45537040 | 10 | 72059526 | *ASCC1* | 3.54 | 1.37e-04 | 4.39e-02 | 1.28 | 4.74e-02 |
| rs77310736 | 7 | 2652743 | *IQCE* | 0.80 | 1.37e-04 | 4.39e-02 | 0.71 | 4.39e-03 |

### pQTL reveals 5 unique variants for Hemoglobin zeta subunit

The pQTL analysis was conducted using 2,941 plasma proteins. Associations between 2,643 GWAS genetic variants and protein abundance were first assessed in a discovery set of 181 samples with WGS data and then evaluated in a replication set of 181 samples. Using a joint significance criterion of FDR-adjusted p-value ≤ 0.05 in one dataset and nominal p-value ≤ 0.05 in the other dataset, we validated 5 pQTL-protein associations (**Table 5**), involving 5 unique variants and one protein, Hemoglobin zeta (HBZ) encoded by the gene Hemoglobin subunit zeta (*HBZ*).

**Table 5:** Top replicated cis-pQTLs ranked by significance in the discovery dataset. Each of the variants reached GWAS p-value ≤ 70.0001 in one or more of the TNBC GWAS studies listed in Table 3. For each pQTL, genomic position in the GRCh38 build, target gene/transcript, and effect estimates (β), raw p-values, and FDR-adjusted p-values in the discovery set are shown, along with corresponding β and p-values in the validation set. All associations shown met replication criteria (FDR ≤ 0.05 in discovery and nominal p ≤ 0.05 in validation).

| eQTL | Chr. | Position<br>(Build 38) | Protein | Discovery |  |  | Validation |  |
| --- | --- | --- | --- | --- | --- | --- | --- | --- |
|  |  |  |  | Beta | P | FDR | Beta | P |
| rs13337843 | 16 | 156204 | HBZ | 2.04 | 1.71E-13 | 4.96E-10 | 2.18 | 3.90E-14 |
| rs13331856 | 16 | 156229 | HBZ | 2.04 | 1.71E-13 | 4.96E-10 | 2.18 | 3.38E-14 |
| rs149127794 | 16 | 146167 | HBZ | 2.09 | 1.33E-12 | 2.57E-09 | 2.10 | 1.84E-13 |
| rs112234623 | 16 | 157048 | HBZ | 1.98 | 3.77E-12 | 5.48E-09 | 2.11 | 9.05E-15 |
| rs13336827 | 16 | 143089 | HBZ | 2.02 | 4.11E-11 | 4.77E-08 | 2.38 | 3.40E-15 |

### Concordant HBZ signals across transcriptomic and proteomic layers

Two HBZ transcripts (ENST00000252951 and ENST00000652335) passed QC filters and were retained for the eQTL analysis. However, ENST00000252951 was expressed in most samples (91% of the 242 individuals), whereas ENST00000652335 was detected in fewer than 20% of samples. It is therefore reassuring that the associations we identified involve the most abundantly expressed and biologically relevant transcript.

Across the integrated analyses, we identified five variants that were positively associated with the *HBZ* protein-coding transcript ENST00000252951 and were also significantly associated with circulating HBZ protein levels. This concordance between transcriptomic and proteomic signals highlights a consistent molecular footprint of HBZ regulation and underscores the robustness of these associations across omics layers (**Figure 2** and **Figure 3**). The overlapping variants consisted of four single-nucleotide polymorphisms (SNPs) (rs13337843, rs13336827, rs112234623, and rs13331856), and one insertion/deletion variant (INDEL) (rs149127794). All five variants are common in our study population (MAF 0.06-0.07 in GENE-FORECAST), reinforcing their potential biological relevance and increasing confidence in their detectability in population-based datasets.

**Figure 2:**
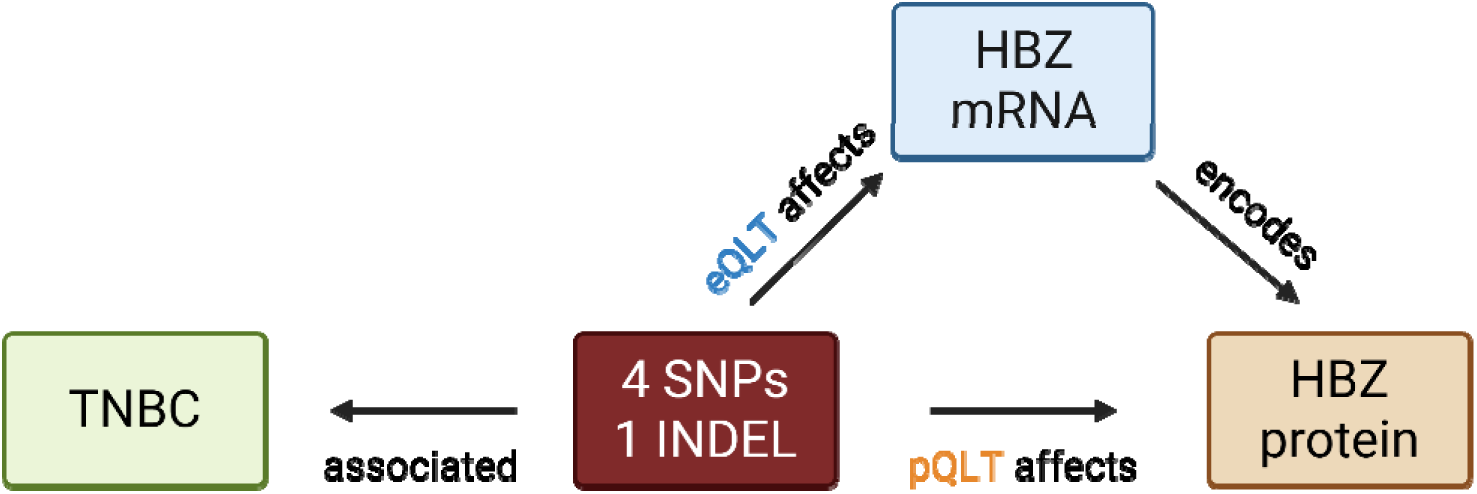
Integrated overview of genetic variants influencing *HBZ* expression across molecular layers. Four SNPs and one INDEL associated with TNBC status were identified as cis-eQTLs for *HBZ* mRNA and cis-pQTLs for circulating HBZ protein levels. The diagram illustrates the concordant regulatory effects of these variants on HBZ transcript and protein abundance, supporting a shared genetic architecture linking genomic variation, *HBZ* expression, and TNBC-related biology.

**Figure 3:**
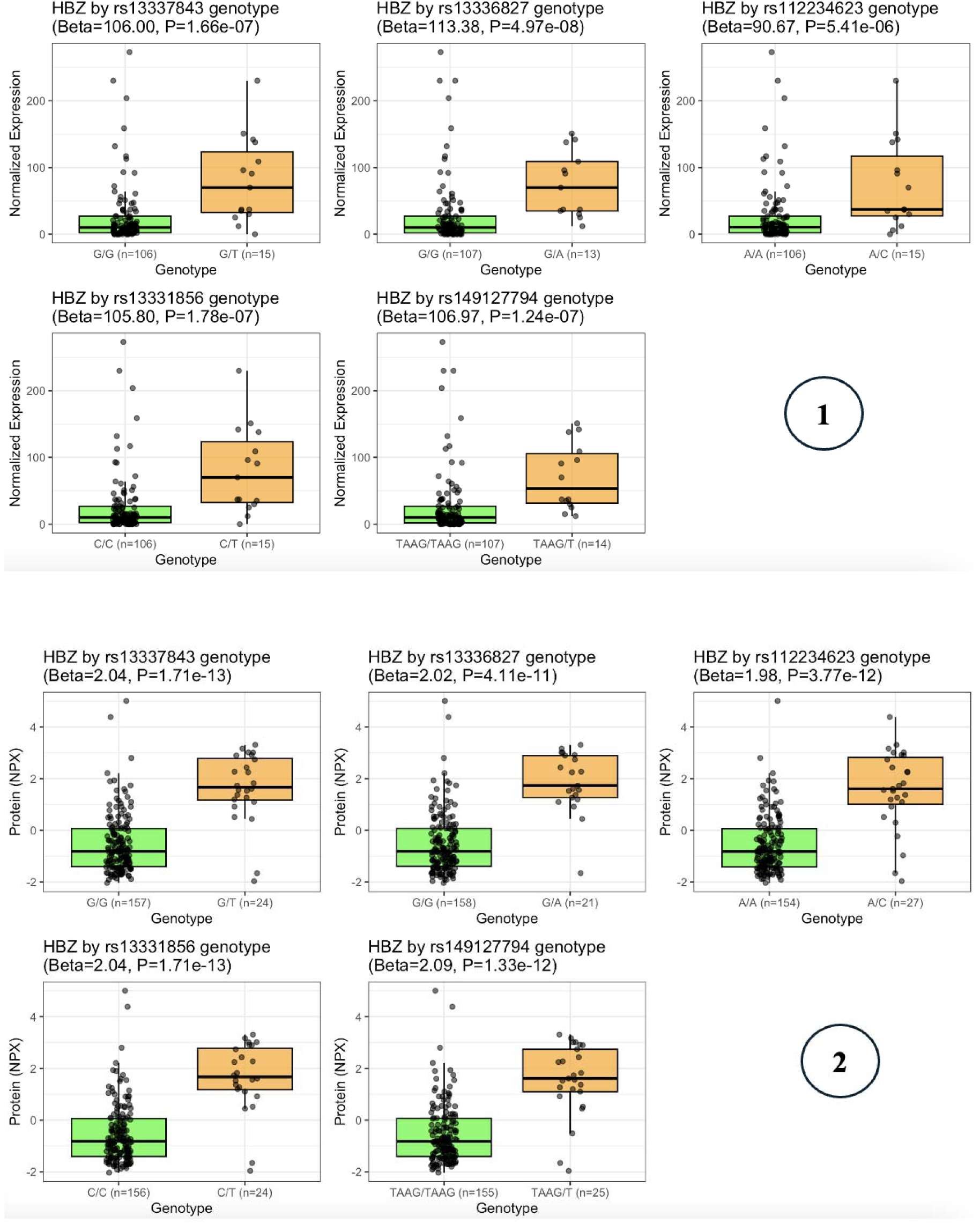
Genotype-Dependent Upregulation of HBZ mRNA and Protein. Boxplots showing the association of the five identified variants with (Panel 1) *HBZ* mRNA expression (normalized counts) and (Panel 2) HBZ plasma protein levels (NPX units) in the discovery dataset. (x-axis: Genotypes (Reference Homozygote vs. Heterozygote). y-axis: Expression levels). For all five variants, carriers of the risk allele (heterozygotes) exhibit significantly higher levels of HBZ transcript and protein compared to non-carriers. No individuals homozygous for the risk allele were observed, suggesting the variant is rare or potentially under strong selection.

All five variants are in strong linkage disequilibrium, as illustrated in **Figure 3**, computed from the GENE-FORECAST data.

**Table 6** summarizes the five HBZ-associated variants identified in the integrative eQTL–pQTL analysis, reporting their minor (A1) and major (A2) alleles, MAF in the GENE-FORECAST cohort, genomic location relative to the HBZ gene, and corresponding odds ratios and p-values from the TNBC GWAS conducted by Jia et al ^12^. The GWAS results from Jia et al. indicate that four of the five HBZ-linked variants are associated with increased TNBC risk, whereas rs112234623 shows a protective association (OR = 0.80).

**Table 6:**
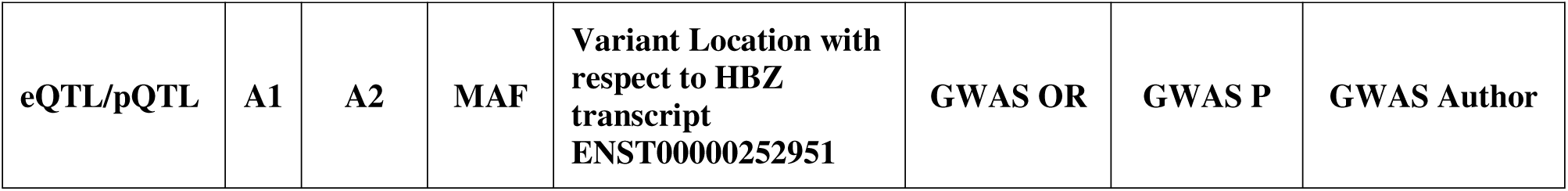
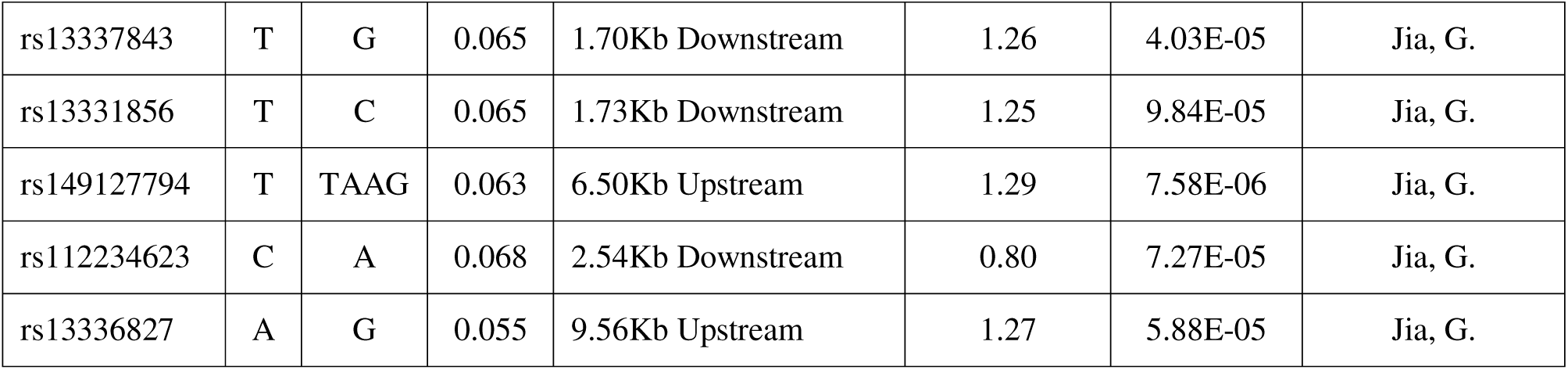
MAF in our study data and GWAS associations for HBZ-linked eQTL/pQTL variants.

### HBZ-associated variants are specific to populations of African ancestry

TNBC is disproportionately prevalent among women of African ancestry, making it important to determine whether the genetic variants identified in our analysis also show ancestry-specific frequency patterns. To address this, we examined allele frequencies for the four SNPs and one INDEL across our GENE-FORECAST cohort and 3 larger human genetics studies, the 1000 Genomes Project, BioVU, and the All of Us Research Program.

Across all datasets, the same pattern emerged: the five HBZ-associated variants are consistently observed at appreciable frequencies only in individuals of African or admixed African ancestry, while being extremely rare or completely absent in European, Asian, and most other global populations (**Table 7**). African and African American groups in GENE-FORECAST, 1000 Genomes, BioVU, and All of Us all showed similar minor allele frequencies, whereas non-African groups displayed near-zero frequencies across all variants.

**Table 7:**
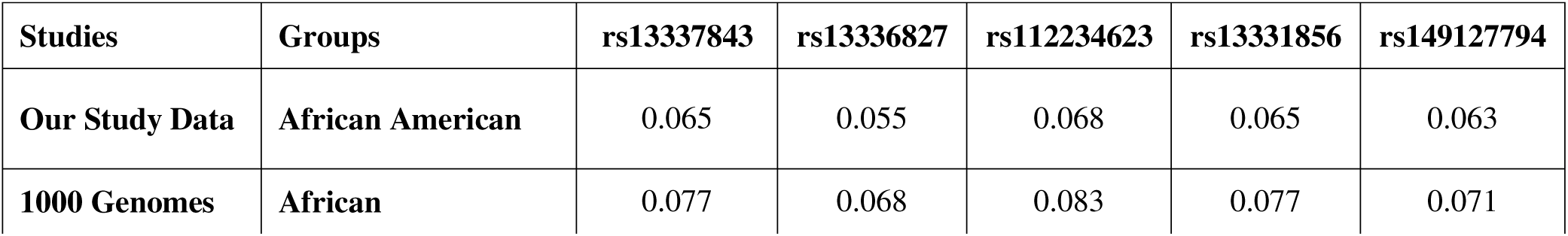
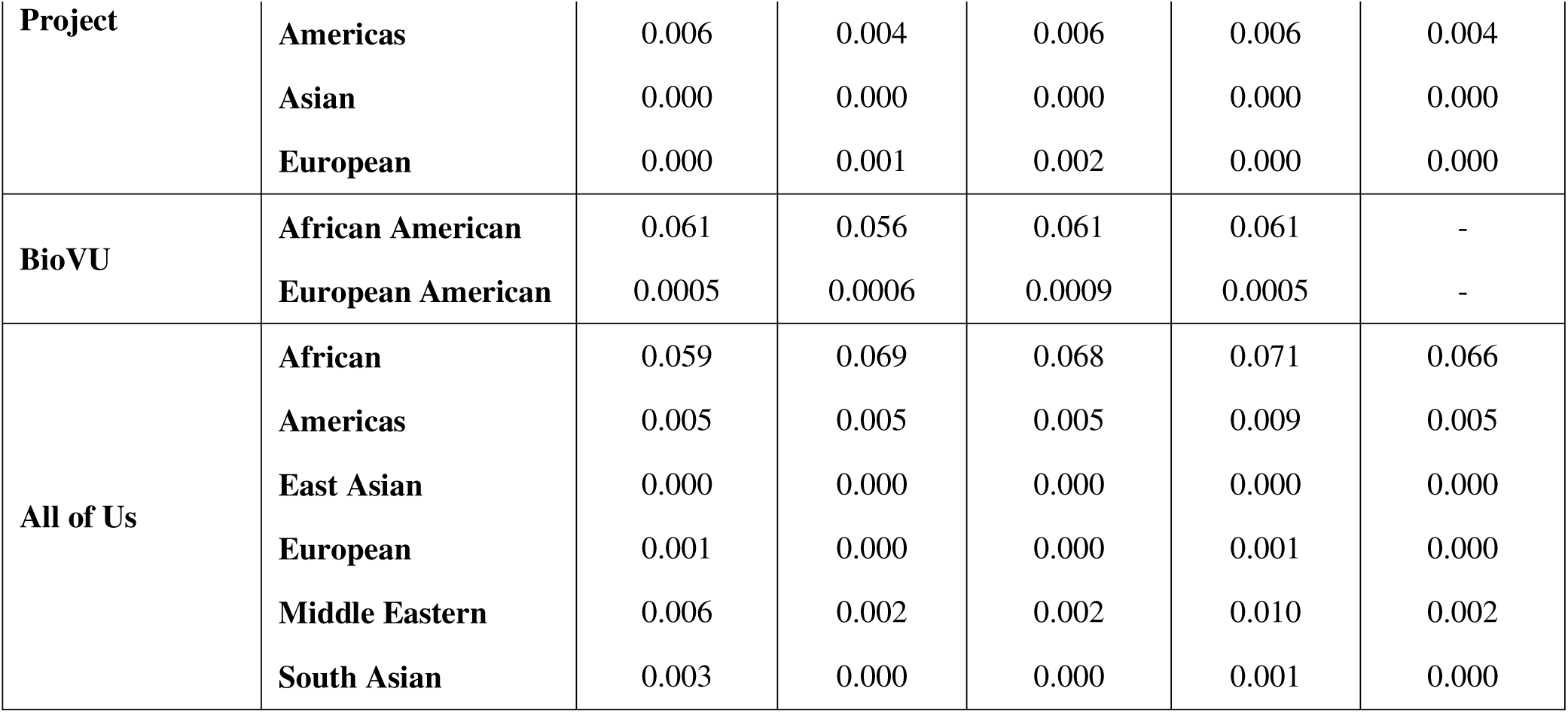
Allele frequencies of the five HBZ-associated variants across multiple datasets show enrichment in African and admixed African populations.

This cross-repository consistency indicates that the HBZ-linked variants form an ancestry-specific signal enriched in African-ancestry populations, aligning with the higher burden of TNBC in these groups and reinforcing the relevance of these findings for understanding population-specific breast cancer risk.

### External validation of eQTLs in GTEx and absence of alternate-allele homozygotes

To further contextualize these associations, we assessed whether the HBZ-linked variants identified in our analysis had been previously reported as eQTLs in the Genotype-Tissue Expression (GTEx) project ^23^. GTEx is the largest and most comprehensive resource for human cis-eQTL mapping, and whole blood, the tissues used in our transcriptomic analysis, is among the well-powered tissues in the GTEx consortium. Four of the five variants (rs13337843, rs112234623, rs13331856, rs149127794) were present in GTEx whole blood, and all four showed significant associations with HBZ expression, with effect directions consistent with those observed in our dataset. This concordance across two independent whole-blood datasets strengthens the robustness of the regulatory signal.

Notably, no participants in our study were homozygous for the alternate allele for any of the variants. The absence of alternate-allele homozygotes, despite allele frequencies in the 6-12% range, raises the intriguing possibility that carrying two copies of the variant allele may have an even more pronounced or potentially distinct impact on *HBZ* regulation, downstream biology, and its impact on the etiology of TNBC.

### Functional Fine-Mapping Identifies rs13336827 as a Putative Driver

To infer the potential causal variant among the five linked polymorphisms, we performed *in silico* fine-mapping using HaploReg v4.1 ^24^. This analysis prioritized rs13336827 as the most likely functional driver.

Unlike the other linked variants, rs13336827 maps to a verified active enhancer region specifically in breast tissue (BRST), as well as blood and liver. It is located within the *NPRL3* gene, which houses the upstream regulatory elements known to control the alpha-globin gene cluster. Functional annotation revealed that this variant acts as a high-occupancy regulatory hub, binding 19 transcription factors. Crucially, this list includes CTCF, a chromatin architectural protein that mediates looping, and GATA1, the master transcriptional regulator of globin genes. This suggests that the risk allele functions by modulating CTCF-mediated chromatin looping, bringing this breast-specific enhancer into direct physical proximity with the *HBZ* promoter to bypass canonical silencing.

**Figure 4:**
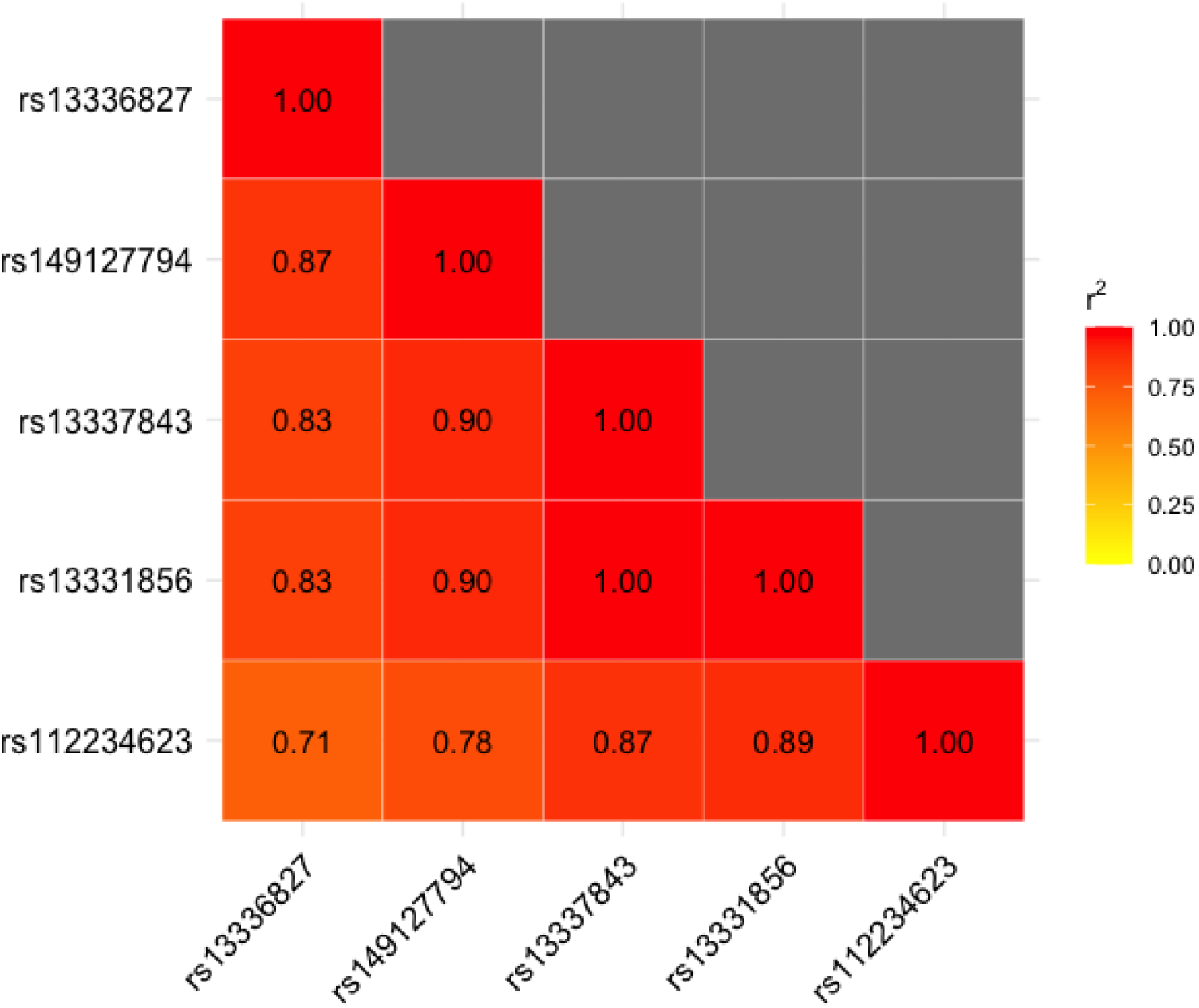
Linkage Disequilibrium (LD) Structure of HBZ Variants. Heatmap displaying the pairwise correlation (r²) between the five identified functional variants. Red squares indicate high LD (r² > 0.8), showing that these variants are strongly correlated and likely inherited together as a single haplotype block in the African American population. This strong correlation implies that these five variants represent a single genetic signal driving *HBZ* expression, rather than five independent effects.

## DISCUSSION

The goal of this integrative, unbiased screening of 2,643 TNBC-associated GWAS variants was to identify novel genes that could explain TNBC disparities. We uncovered a striking and unexpected convergence on Hemoglobin Zeta (HBZ) as the primary functional target supported across both transcriptomic and proteomic layers. Despite beginning with thousands of variants and ∼130,000 mRNA transcripts of ∼16,000 protein-coding genes, only five variants, four SNPs and one INDEL, showed consistent, replicated cis-regulatory effects on HBZ mRNA expression and circulating HBZ protein levels. This cross-layer convergence, demonstrated through a discovery-validation framework within the GENE-FORECAST cohort and further supported by eQTL evidence in GTEx whole blood, identifies HBZ as a previously unrecognized, ancestry-enriched molecular candidate relevant to TNBC biology.

HBZ is an embryonic globin gene silenced shortly after gestation ^25^. Its normal function is to produce the zeta-globin (ζ) chain, an “alpha-like” component of the first embryonic hemoglobins (e.g., Hb Gower 1, ζ□ε□) ^25^. This gene is completely silenced during the hemoglobin switching process that occurs shortly after gestation^18^. The developmental inactivity of HBZ makes its apparent genetic regulation in adults biologically unexpected. TNBC tumors are characterized by rapid proliferation, disrupted vasculature, and profound hypoxia, stressors known to drive metabolic adaptation and tumor progression. HBZ is surprisingly expressed in tissues such as the testis, bone marrow, spleen, and the choroid plexus in the brain, which are also prone to hypoxia. The zeta-globin protein encoded by HBZ forms hemoglobin complexes with extremely high oxygen affinity, enabling efficient oxygen scavenging in these low-oxygen environments. We therefore propose that genetically driven upregulation of HBZ may reflect a reactivation of embryonic oxygen-binding programs that help tumor cells adapt to severe hypoxia. Although this proposed mechanism requires additional experimental validation, it is well grounded in hemoglobin physiology and consistent with the aggressiveness of TNBC that is driven at least in part by hypoxia. Indeed, prior studies in colorectal cancer have demonstrated that aggressive Polyploid Giant Cancer Cells (PGCCs) reactivate embryonic hemoglobin zeta (HBZ) to efficiently bind oxygen and survive hypoxic stress ^26^. This provides a direct precedent for the co-option of this embryonic globin gene by solid tumors as a survival mechanism against metabolic stress.

A key and clinically relevant dimension of our findings is the ancestry-specificity of the HBZ-linked variants. Across multiple population datasets, including GENE-FORECAST, 1000 Genomes, BioVU, and All of Us, the five regulatory variants were consistently present at appreciable frequencies only in African and African-admixed groups, while being nearly absent in Europeans and Asians. This pattern mirrors the disproportionate burden of TNBC in women of African ancestry ^5–7^ and highlights how population-specific genetic architectures may shape disease susceptibility. Importantly, such variants would be undetectable in European-centric genomic studies. Our work, therefore, illustrates how analyses conducted in African ancestry cohorts can reveal biologically important signals that remain hidden when research is limited to homogeneous populations.

The robustness of the HBZ signal is increased by several layers of evidence. The cis-eQTL and cis-pQTL associations were reproduced across discovery and validation subsets of the same cohort, using identical pipelines and covariate structures. The strong linkage disequilibrium among the five variants indicates a coherent regulatory region rather than multiple unrelated signals. Furthermore, four of the five variants appear as significant HBZ eQTLs in GTEx whole blood, an external dataset, with fully concordant effect directions. Finally, the complete absence of alternate-allele homozygotes in our cohort, despite minor allele frequencies in the 6-7% range, suggests possible dosage constraints or selective pressures, warranting further investigation.

All five variants are in strong LD and show a consistent effect of increasing HBZ mRNA and protein abundance in whole blood. Importantly, the HBZ mRNA upregulation we observe is also supported by independent eQTL evidence from GTEx in the same tissue. However, while four of the variants are associated with an increased risk (OR > 1) of TNBC, the rs112234623 variant is reported with a protective effect (OR < 1) in the source GWAS study. The observation that rs112234623 is protective despite being in strong LD with four TNBC risk–increasing variants suggests potential allelic and functional heterogeneity within the HBZ regulatory region. Such patterns are well documented in complex traits, where tightly linked variants exert distinct or even opposing regulatory effects on nearby genes ^16, 19^. Possible explanations include multiple causal variants within the same enhancer, haplotype-specific effects arising from subtle sub-haplotype structure, or variant-specific disruption of transcription factor motifs, as described in prior fine-mapping studies ^27, 28^. It is also conceivable that the protective signal reflects synthetic association or tagging of a different causal allele, a phenomenon reported in LD-dense regions ^29^. Disentangling these mechanisms would require functional dissection and haplotype-level analyses as important next steps, but beyond the scope of the present study.

Although HBZ emerged as the strongest molecular candidate, additional validated cis-eQTL associations were observed. Genes such as SLC27A1, POSTN, SNRNP25, and C11orf54 showed replicated transcript-level associations, some of which align with known TNBC biology, including lipid metabolism, extracellular matrix remodeling, and RNA processing ^30, 31^. However, unlike HBZ, these genes did not show replicated effects at the protein level nor ancestry-specific allele frequency patterns. Thus, while they may represent secondary contributors or context-specific pathways, HBZ remains the only target supported by convergent transcriptomic, proteomic, external eQTL, and population-genetic evidence. Importantly, the Olink Explore 3072 platform measures only a subset of the proteome (2,941 proteins in our dataset), and several of these eQTL-target genes may simply not have had corresponding proteins assayed. Therefore, the absence of replicated pQTLs for these genes likely reflects limited proteomic coverage rather than a definitive lack of biological relevance.

This study has several strengths. It leverages multi-omics data collected in African Americans, a population disproportionately affected by TNBC yet historically underrepresented in cancer genomic research. The unbiased analytical design, starting from GWAS loci and allowing the multi-omics data to identify functional targets, minimizes prior assumptions and enables the discovery of an unexpected embryonic gene. Critically, both the transcriptomic and proteomic data utilized whole blood as the source tissue, which provides a systemic view of molecular perturbation linking the TNBC variants to HBZ expression. The integration of WGS, gene expression, and proteomics within the same individuals, combined with a discovery-validation structure, enhances the internal reproducibility of our findings.

We also acknowledge limitations. Gene expression and protein measurements were conducted in whole blood, not tumor tissue, which may not fully capture transcriptional activity in breast epithelial or tumor cells. The sample size, particularly for eQTL discovery, is modest relative to large consortia. Additionally, the study remains associative; mechanistic experiments are needed to determine whether HBZ reactivation influences TNBC initiation, progression, or therapeutic vulnerability. Our future work will build directly on these findings through (i) biological validation of HBZ overexpression in TNBC tumors, particularly among patients of African ancestry, (ii) functional perturbation experiments using siRNA or CRISPR knockout of HBZ in African-ancestry TNBC cell lines to assess its role in hypoxia-driven survival, and (iii) mechanistic assays, such as luciferase reporter systems, to determine whether the five regulatory variants directly modulate HBZ transcription. These planned experiments are outlined in detail in the Future Directions section.

### Conclusions

In summary, our integrative multi-omics analysis identifies HBZ as a high-confidence, ancestry-enriched molecular candidate regulated by TNBC-associated genetic variants. Although the effect sizes (ORs) of these variants are modest, this is entirely consistent with the polygenic architecture of TNBC and most common cancers, where individual variants typically exert small but biologically meaningful effects. Modest ORs do not preclude functional relevance; rather, they often reflect fine-tuned regulatory perturbations that cumulatively shift gene expression programs and disease susceptibility, particularly when they converge on a shared pathway, as observed here with HBZ regulation.

Our findings suggest a plausible biological model, reactivation of an embryonic oxygen-scavenging program, that may help explain both the aggressive behavior of TNBC and its disproportionate impact among women of African ancestry. More broadly, this work underscores the necessity of including diverse populations in genomic research to uncover ancestry-specific regulatory mechanisms, improve equity in cancer biology, and ensure that molecular insights translate into benefits for all populations.

## Data Availability

All data produced in the present study are available upon reasonable request to the authors

## REFERENCES

1. Dogra AK, Prakash A, Gupta S, Gupta M. Prognostic significance and molecular classification of triple negative breast cancer: A systematic review. Eur J Breast Health. 2025;21(2):101–14. doi: 10.4274/ejbh.galenos.2025.2024-10-2; PMCID: PMC11934825.

2. Society AC. Treatment of triple-negative breast cancer 2022.

3. Liedtke C, Mazouni C, Hess KR, André F, Tordai A, Mejia JA, Symmans WF, Gonzalez-Angulo AM, Hennessy B, Green M, Cristofanilli M, Hortobagyi GN, Pusztai L. Response to neoadjuvant therapy and long-term survival in patients with triple-negative breast cancer. J Clin Oncol. 2008;26(8):1275–81. doi: 10.1200/JCO.2007.14.4147.

4. Srivastava N, Usmani SS, Subbarayan R, Saini R, Pandey PK. Hypoxia: syndicating triple negative breast cancer against various therapeutic regimens. Front Oncol. 2023;13:1199105. doi: 10.3389/fonc.2023.1199105; PMCID: PMC10363988.

5. Stark A, Kleer CG, Martin I, Awuah B, Nsiah-Asare A, Takyi V, Braman M, Quayson SE, Zarbo R, Wicha M, Newman L. African ancestry and higher prevalence of triple-negative breast cancer: findings from an international study. Cancer. 2010;116(21):4926–32. doi: 10.1002/cncr.25276; PMCID: PMC3138711.

6. Sung H, Wiese D, Jatoi I, Jemal A. State variation in racial and ethnic disparities in incidence of triple-negative breast cancer among US women. JAMA Oncol. 2023;9(5):700–4. doi: 10.1001/jamaoncol.2022.7835; PMCID: PMC9982739.

7. Hercules SM, Alnajar M, Chen C, Mladjenovic SM, Shipeolu BA, Perkovic O, Pond GR, Mbuagbaw L, Blenman KR, Daniel JM. Triple-negative breast cancer prevalence in Africa: a systematic review and meta-analysis. BMJ Open. 2022;12(5):e055735. Epub 20220527. doi: 10.1136/bmjopen-2021-055735. PubMed PMID: 35623750; PMCID: PMC9150263.

8. Buniello A, MacArthur JAL, Cerezo M, Harris LW, Hayhurst J, Malangone C, McMahon A, Morales J, Mountjoy E, Sollis E, Suveges D, Vrousgou O, Whetzel PL, Amode R, Guillen JA, Riat HS, Trevanion SJ, Hall P, Junkins H, Flicek P, Burdett T, Hindorff LA, Cunningham F, Parkinson H. The NHGRI-EBI GWAS Catalog of published genome-wide association studies, targeted arrays and summary statistics 2019. Nucleic Acids Res. 2019;47(D1):D1005–D12. doi: 10.1093/nar/gky1120; PMCID: PMC6323933.

9. Zhang H, Ahearn TU, Lecarpentier J, Barnes D, Beesley J, Qi G, Jiang X, O’Mara TA, Zhao N, Bolla MK, Dunning AM, Dennis J, Wang Q, Ful ZA, Aittomäki K, Andrulis IL, Anton-Culver H, Arndt V, Aronson KJ, Arun BK, Auer PL, Azzollini J, Barrowdale D, Becher H, Beckmann MW, Behrens S, Benitez J, Bermisheva M, Bialkowska K, Blanco A, Blomqvist C, Bogdanova NV, Bojesen SE, Bonanni B, Bondavalli D, Borg A, Brauch H, Brenner H, Briceno I, Broeks A, Brucker SY, Brüning T, Burwinkel B, Buys SS, Byers H, Caldés T, Caligo MA, Calvello M, Campa D, Castelao JE, Chang-Claude J, Chanock SJ, Christiaens M, Christiansen H, Chung WK, Claes KBM, Clarke CL, Cornelissen S, Couch FJ, Cox A, Cross SS, Czene K, Daly MB, Devilee P, Diez O, Domchek SM, Dörk T, Dwek M, Eccles DM, Ekici AB, Evans DG, Fasching PA, Figueroa J, Foretova L, Fostira F, Friedman E, Frost D, Gago-Dominguez M, Gapstur SM, Garber J, García-Sáenz JA, Gaudet MM, Gayther SA, Giles GG, Godwin AK, Goldberg MS, Goldgar DE, González-Neira A, Greene MH, Gronwald J, Guénel P, Häberle L, Hahnen E, Haiman CA, Hake CR, Hall P, Hamann U, Harkness EF, Heemskerk-Gerritsen BAM, Hillemanns P, Hogervorst FBL, Holleczek B, Hollestelle A, Hooning MJ, Hoover RN, Hopper JL, Howell A, Huebner H, Hulick PJ, Imyanitov EN, kConFab I, Investigators A, Isaacs C, Izatt L, Jager A, Jakimovska M, Jakubowska A, James P, Janavicius R, Janni W, John EM, Jones ME, Jung A, Kaaks R, Kapoor PM, Karlan BY, Keeman R, Khan S, Khusnutdinova E, Kitahara CM, Ko Y-D, Konstantopoulou I, Koppert LB, Koutros S, Kristensen VN, Laenkholm A-V, Lambrechts D, Larsson SC, Laurent-Puig P, Lazaro C, Lazarova E, Lejbkowicz F, Leslie G, Lesueur F, Lindblom A, Lissowska J, Lo W-Y, Loud JT, Lubinski J, Lukomska A, MacInnis RJ, Mannermaa A, Manoochehri M, Manoukian S, Margolin S, Martinez ME, Matricardi L, McGuffog L, McLean C, Mebirouk N, Meindl A, Menon U, Miller A, Mingazheva E, Montagna M, Mulligan AM, Mulot C, Muranen TA, Nathanson KL, Neuhausen SL, Nevanlinna H, Neven P, Newman WG, Nielsen FC, Nikitina-Zake L, Nodora J, Offit K, Olah E, Olopade OI, Olsson H, Orr N, Papi L, Papp J, Park-Simon T-W, Parsons MT, Peissel B, Peixoto A, Peshkin B, Peterlongo P, Peto J, Phillips K-A, Piedmonte M, Plaseska-Karanfilska D, Prajzendanc K, Prentice R, Prokofyeva D, Rack B, Radice P, Ramus SJ, Rantala J, Rashid MU, Rennert G, Rennert HS, Risch HA, Romero A, Rookus MA, Rübner M, Rüdiger T, Saloustros E, Sampson S, Sandler DP, Sawyer EJ, Scheuner MT, Schmutzler RK, Schneeweiss A, Schoemaker MJ, Schöttker B, Schürmann P, Senter L, Sharma P, Sherman ME, Shu X-O, Singer CF, Smichkoska S, Soucy P, Southey MC, Spinelli JJ, Stone J, Stoppa-Lyonnet D, Study E, Collaborators GS, Swerdlow AJ, Szabo CI, Tamimi RM, Tapper WJ, Taylor JA, Teixeira MR, Terry M, Thomassen M, Thull DL, Tischkowitz M, Toland AE, Tollenaar RAEM, Tomlinson I, Torres D, Troester MA, Truong T, Tung N, Untch M, Vachon CM, van den Ouweland AMW, van der Kolk LE, van Veen EM, vanRensburg EJ, Vega A, Wappenschmidt B, Weinberg CR, Weitzel JN, Wildiers H, Winqvist R, Wolk A, Yang XR, Yannoukakos D, Zheng W, Zorn KK, Milne RL, Kraft P, Simard J, Pharoah PDP, Michailidou K, Antoniou AC, Schmidt MK, Chenevix-Trench G, Easton DF, Chatterjee N, García-Closas M. Genome-wide association study identifies 32 novel breast cancer susceptibility loci from overall and subtype-specific analyses. Nat Genet. 2020;52(6):572–81. doi: 10.1038/s41588-020-0609-2; PMCID: PMC7808397.

10. Purrington KS, Slager S, Eccles D, Yannoukakos D, Fasching PA, Miron P, Carpenter J, Chang-Claude J, Martin NG, Montgomery GW, Kristensen V, Anton-Culver H, Goodfellow P, Tapper WJ, Rafiq S, Gerty SM, Durcan L, Konstantopoulou I, Fostira F, Vratimos A, Apostolou P, Konstanta I, Kotoula V, Lakis S, Dimopoulos MA, Skarlos D, Pectasides D, Fountzilas G, Beckmann MW, Hein A, Ruebner M, Ekici AB, Hartmann A, Schulz-Wendtland R, Renner SP, Janni W, Rack B, Scholz C, Neugebauer J, Andergassen U, Lux MP, Haeberle L, Clarke C, Pathmanathan N, Rudolph A, Flesch-Janys D, Nickels S, Olson JE, Ingle JN, Olswold C, Slettedahl S, Eckel-Passow JE, Anderson SK, Visscher DW, Cafourek VL, Sicotte H, Prodduturi N, Weiderpass E, Bernstein L, Ziogas A, Ivanovich J, Giles GG, Baglietto L, Southey M, Kosma V-M, Fischer H-P, Network G, Reed MWR, Cross SS, Deming-Halverson S, Shrubsole M, Cai Q, Shu X-O, Daly M, Weaver J, Ross E, Klemp J, Sharma P, Torres D, Rüdiger T, Wölfing H, Ulmer H-U, Försti A, Khoury T, Kumar S, Pilarski R, Shapiro CL, Greco D, Heikkilä P, Aittomäki K, Blomqvist C, Irwanto A, Liu J, Pankratz VS, Wang X, Severi G, Mannermaa A, Easton D, Hall P, Brauch H, Cox A, Zheng W, Godwin AK, Hamann U, Ambrosone C, Toland AE, Nevanlinna H, Vachon CM, Couch FJ. Genome-wide association study identifies 25 known breast cancer susceptibility loci as risk factors for triple-negative breast cancer. Carcinogenesis. 2014;35(5):1012–9. doi: 10.1093/carcin/bgt404; PMCID: PMC4004200.

11. Sun X, Verma SP, Jia G, Wang X, Ping J, Guo X, Shu X-O, Chen J, Derkach A, Cai Q, Liang X, Long J, Offit K, Oh JH, Reiner AS, Watt GP, Woods M, Yang Y, Ambrosone CB, Ambs S, Chen Y, Concannon P, Garcia-Closas M, Gu J, Haiman CA, Hu JJ, Huo D, John EM, Knight JA, Li CI, Lynch CF, Mellemkjær L, Nathanson KL, Nemesure B, Olopade OI, Olshan AF, Pal T, Palmer JR, Press MF, Sanderson M, Sandler DP, Troester MA, Zheng W, Bernstein JL, Buas MF, Shu X. Case-case genome-wide analyses identify subtype-informative variants that confer risk for breast cancer. Cancer Res. 2024;84(15):2533–48. doi: 10.1158/0008-5472.CAN-23-3854; PMCID: PMC11293972.

12. Jia G, Ping J, Guo X, Yang Y, Tao R, Li B, Ambs S, Barnard ME, Chen Y, Garcia-Closas M, Gu J, Hu JJ, Huo D, John EM, Li CI, Li JL, Nathanson KL, Nemesure B, Olopade OI, Pal T, Press MF, Sanderson M, Sandler DP, Shu X-O, Troester MA, Yao S, Adejumo PO, Ahearn T, Brewster AM, Hennis AJM, Makumbi T, Ndom P, O’Brien KM, Olshan AF, Oluwasanu MM, Reid S, Butler EN, Huang M, Ntekim A, Qian H, Zhang H, Ambrosone CB, Cai Q, Long J, Palmer JR, Haiman CA, Zheng W. Genome-wide association analyses of breast cancer in women of African ancestry identify new susceptibility loci and improve risk prediction. Nat Genet. 2024;56(5):819–26. doi: 10.1038/s41588-024-01736-4; PMCID: PMC11284829.

13. Abbas M, Martin P, Lindsey ML, Bennett ES, Brown TL, Nzerue C, Williams CR, Gaye A. Intersecting transcriptomic landscapes of hypertension and kidney function in African American women. Am J Physiol Renal Physiol. 2025;329(1):F59–F70. Epub 20250530. doi: 10.1152/ajprenal.00067.2025. PubMed PMID: 40445960; PMCID: PMC12316382.

14. Institute B. broadinstitute/gtex-pipeline GitHub 2015 [cited 2018]. Available from: https://github.com/broadinstitute/gtex-pipeline.

15. Robinson MD, Oshlack A. A scaling normalization method for differential expression analysis of RNA-seq data. Genome Biol. 2010;11(3):R25. Epub 2010/03/04. doi: 10.1186/gb-2010-11-3-r25. PubMed PMID: 20196867; PMCID: PMC2864565.

16. Farh KK, Marson A, Zhu J, Kleinewietfeld M, Housley WJ, Beik S, Shoresh N, Whitton H, Ryan RJ, Shishkin AA, Hatan M, Carrasco-Alfonso MJ, Mayer D, Luckey CJ, Patsopoulos NA, De Jager PL, Kuchroo VK, Epstein CB, Daly MJ, Hafler DA, Bernstein BE. Genetic and epigenetic fine mapping of causal autoimmune disease variants. Nature. 2015;518(7539):337–43. Epub 20141029. doi: 10.1038/nature13835. PubMed PMID: 25363779; PMCID: PMC4336207.

17. Trynka G, Sandor C, Han B, Xu H, Stranger BE, Liu XS, Raychaudhuri S. Chromatin marks identify critical cell types for fine mapping complex trait variants. Nat Genet. 2013;45(2):124–30. Epub 20121223. doi: 10.1038/ng.2504. PubMed PMID: 23263488; PMCID: PMC3826950.

18. Maurano MT, Humbert R, Rynes E, Thurman RE, Haugen E, Wang H, Reynolds AP, Sandstrom R, Qu H, Brody J, Shafer A, Neri F, Lee K, Kutyavin T, Stehling-Sun S, Johnson AK, Canfield TK, Giste E, Diegel M, Bates D, Hansen RS, Neph S, Sabo PJ, Heimfeld S, Raubitschek A, Ziegler S, Cotsapas C, Sotoodehnia N, Glass I, Sunyaev SR, Kaul R, Stamatoyannopoulos JA. Systematic localization of common disease-associated variation in regulatory DNA. Science. 2012;337(6099):1190–5. Epub 20120905. doi: 10.1126/science.1222794. PubMed PMID: 22955828; PMCID: PMC3771521.

19. Consortium GT, Laboratory DA, Coordinating Center - Analysis Working G, Statistical Methods groups-Analysis Working G, Enhancing Gg, Fund NIHC, Nih/Nci, Nih/Nhgri, Nih/Nimh, Nih/Nida, Biospecimen Collection Source Site N, Biospecimen Collection Source Site R, Biospecimen Core Resource V, Brain Bank Repository-University of Miami Brain Endowment B, Leidos Biomedical-Project M, Study E, Genome Browser Data I, Visualization EBI, Genome Browser Data I, Visualization-Ucsc Genomics Institute UoCSC, Lead a, Laboratory DA, Coordinating C, management NIHp, Biospecimen c, Pathology, e QTLmwg, Battle A, Brown CD, Engelhardt BE, Montgomery SB. Genetic effects on gene expression across human tissues. Nature. 2017;550(7675):204–13. doi: 10.1038/nature24277. PubMed PMID: 29022597; PMCID: PMC5776756.

20. Shabalin AA. Matrix eQTL: ultra fast eQTL analysis via large matrix operations. Bioinformatics. 2012;28(10):1353–8. Epub 2012/04/12. doi: 10.1093/bioinformatics/bts163. PubMed PMID: 22492648; PMCID: PMC3348564.

21. Fentiman I. Male breast cancer: a review. Ecancermedicalscience. 2009;3:140. Epub 20090320. doi: 10.3332/ecancer.2009.140. PubMed PMID: 22276005; PMCID: PMC3223984.

22. NIH. Consideration of Sex as a Biological Variable in NIH-funded Research 2015. Available from: https://grants.nih.gov/grants/guide/notice-files/not-od-15-102.html.

23. Consortium GT. The Genotype-Tissue Expression (GTEx) project. Nat Genet. 2013;45(6):580–5. doi: 10.1038/ng.2653. PubMed PMID: 23715323; PMCID: PMC4010069.

24. Ward LD, Kellis M. HaploReg: a resource for exploring chromatin states, conservation, and regulatory motif alterations within sets of genetically linked variants. Nucleic Acids Res. 2012;40(Database issue):D930–4. Epub 20111107. doi: 10.1093/nar/gkr917. PubMed PMID: 22064851; PMCID: PMC3245002.

25. Stamatoyannopoulos G. Control of globin gene expression during development and erythroid differentiation. Exp Hematol. 2005;33(3):259–71. doi: 10.1016/j.exphem.2004.11.007; PMCID: PMC2819985.

26. Li Z, Zheng M, Zhang H, Yang X, Fan L, Fu F, Fu J, Niu R, Yan M, Zhang S. Arsenic Trioxide Promotes Tumor Progression by Inducing the Formation of PGCCs and Embryonic Hemoglobin in Colon Cancer Cells. Front Oncol. 2021;11:720814. Epub 20211005. doi: 10.3389/fonc.2021.720814. PubMed PMID: 34676163; PMCID: PMC8523995.

27. Claussnitzer M, Dankel SN, Kim KH, Quon G, Meuleman W, Haugen C, Glunk V, Sousa IS, Beaudry JL, Puviindran V, Abdennur NA, Liu J, Svensson PA, Hsu YH, Drucker DJ, Mellgren G, Hui CC, Hauner H, Kellis M. FTO Obesity Variant Circuitry and Adipocyte Browning in Humans. N Engl J Med. 2015;373(10):895–907. Epub 20150819. doi: 10.1056/NEJMoa1502214. PubMed PMID: 26287746; PMCID: PMC4959911.

28. Corradin O, Scacheri PC. Enhancer variants: evaluating functions in common disease. Genome Med. 2014;6(10):85. Epub 20141028. doi: 10.1186/s13073-014-0085-3. PubMed PMID: 25473424; PMCID: PMC4254432.

29. Dickson SP, Wang K, Krantz I, Hakonarson H, Goldstein DB. Rare variants create synthetic genome-wide associations. PLoS Biol. 2010;8(1):e1000294. Epub 20100126. doi: 10.1371/journal.pbio.1000294. PubMed PMID: 20126254; PMCID: PMC2811148.

30. Mendes C, Lopes-Coelho F, Ramos C, Martins F, Santos I, Rodrigues A, Silva F, André S, Serpa J. Unraveling FATP1, regulated by ER-β, as a targeted breast cancer innovative therapy. Sci Rep. 2019;9(1):14107. doi: 10.1038/s41598-019-50531-3; PMCID: PMC6773857.

31. Moreira-Barbosa C, Matos A, Fernandes R, Mendes-Ferreira M, Rodrigues R, Cruz T, Costa ÂM, Cardoso AP, Ghilardi C, Oliveira MJ, Ribeiro R. The role of fatty acids metabolism on cancer progression and therapeutics development. Bioactive Lipids: Elsevier; 2023. p. 101–32.

